# Time scale performance of rapid antigen testing for SARS-COV-2: evaluation of ten rapid antigen assays

**DOI:** 10.1101/2021.05.27.21257868

**Authors:** Zakarya Abusrewil, Inas M Alhudiri, Hamza Hussein Kaal, Salah Edin El Meshri, Fawzi O. Ebrahim, Tarek Dalyoum, Abdussamee A Efrefer, Khaled Ibrahim, Mohammed Ben Elfghi, Suleiman Abusrewil, Adam Elzagheid

## Abstract

There is a great demand for more rapid tests for SARS-COV-2 detection to reduce waiting time, boost public health strategies for combating disease, decrease costs, and prevent overwhelming laboratory capacities. This study was conducted to assess the performance of 10 lateral flow device viral antigen immunoassays for the detection of SARS-CoV-2 in nasopharyngeal swab specimens. We analyzed 231 nasopharyngeal samples collected from October 2020-December 2020, from suspected COVID-19 cases and contacts of positive cases at Biotechnology Research Center laboratories, Tripoli, Libya. The performance of 10 COVID-19 Antigen (Ag) rapid test devices for the detection of SARS-CoV-2 antigen was compared to RT-qPCR. In this study 161 cases had symptoms consistent with COVID-19. The mean duration from symptom onset was 6.6 ±4.3 days. The median cycle threshold (Ct) of positive samples was 25. Among the 108 positive samples detected by RT-qPCR, the COVID-19 antigen (Ag) tests detected 83 cases correctly. All rapid Ag test devices used in this study showed 100% specificity. While tests from 6 manufacturers had an overall sensitivity range from 75-100%, the remaining 4 tests had sensitivity of 50-71.43%. Sensitivity during the first 6 days of symptoms and in samples with high viral loads (Ct<25), was 100% in all but 2 of the test platforms. False negative samples had a median Ct of 34 and an average duration of onset of symptoms of 11.3 days (range=5-20 days). Antigen test diagnosis has high sensitivity and specificity in early disease when patients present less than 7 days of symptom onset. Patients are encouraged to test as soon as they get COVID-19 related symptoms within 1 week and to seek medical advice within 24 hrs. if they develop disturbed smell/taste. The use of rapid antigen tests is important for controlling COVID-19 pandemic and reducing burden on molecular diagnostic laboratories.

## Introduction

Nucleic acid amplification test using RT-qPCR assay is currently the mainstay diagnostic test for COVID-19 in most laboratories. However, results of RT-qPCR assays can take several days to deliver, hampering the disease containment efforts. Consequently, there is an emerging demand for more rapid and easier tests to reduce waiting time, boost public health strategies for combating disease, decrease costs, and prevent overwhelming laboratory capacities.

SARS-CoV-2-rapid antigen tests have been largely developed and many countries have adopted them for diagnosis in triage and hospital settings and at points of entry ^1^. However, many of these tests have not been independently validated. The World Health Organization (WHO) encourages laboratories to evaluate the performance of commercial rapid antigen assays to update the current evidence and recommend specific test kits ^2^.

According to previous studies, rapid antigen tests could be used in high-risk populations to quickly identify patients with infection and prevent disease transmission by repeat testing^3^. Evaluation of these tests showed also diagnostic performance in screening mass population^4^.

Libyan health authorities have approved the use of rapid antigen tests in triage, isolation centers and hospitals ^5^. This study was conducted to assess the performance of 10 antigen-based rapid assays for the detection of SARS-CoV-2 in nasopharyngeal swab specimens from suspected COVID-19 cases and contacts.

## Materials and Methods

### Study Population

This was a prospective single-center study. A total of 231 patients with clinical features suggestive of COVID-19 or a history of close contact with COVID-19 positive patient were enrolled in this study from October 10 ^_^ December 31 2020. However, there were financial difficulties to obtain adequate quantities of antigen test device to study larger sample size for each type during the pandemic and hence the number of samples tested was between 15-39 depending on the kit contents. The majority of patients were having symptoms suggestive of COVID-19 (70%). A quarter of participants were asymptomatic and were in contact with COVID-19 positive case. Few patients were asymptomatic but had no history of contact. Duration since symptom onset was variable among the cases with an average of 7 days (Table 1).

**Table 1.** Characteristics of study patients.

| **Characteristics** | **No (%)** |
| --- | --- |
| **Gender** |  |
| Male | 130 (56.2) |
| Female | 101 (43.7) |
| **Age (Mean±SD)** | 40.8±15.23 |
| **Asymptomatic patients with close contact** | 58 (25) |
| **Days since close contact (Mean±SD)** | 7.1±3 |
| **Asymptomatic no contact** | 12 (5) |
| **Symptomatic patients** | 161 (69.6) |
| **Days from symptom onset (Mean±SD)** | 7±6 |

### Antigen Test procedure

We evaluated the performance of 10 rapid antigen tests for SARS-COV-2. These lateral flow tests are a qualitative membrane-based colloidal gold chromatography immunoassay (Fluorecare SARS-CoV-2 spike protein test, Shenzhen Microprofit Biotech Co; ESPLINE SARS-CoV-2, Fujirbio; RapiGen Covid-19 Ag Detection Kit, Biocredit; Abbott Panbio™ COVID-19 Ag Rapid Test; Flowflex™ SARS-CoV-2 Antigen Rapid Test, Acon; Assut Europe antigen testing COVID-19; Coronavirus antigen rapid test cassette Orient Gene; CerTest SARS-CoV-2 one step card test; CerTest Biotech, Bioperfectus SARS-CoV-2 Antigen Rapid Test Kit; Bioperfectus technologies; AMP Rapid Test SARS-CoV-2 Ag; AMP diagnostics). All assays detect SARS-CoV-2 nucleoprotein except Fluorecare assay which detects spike protein. Tests were performed according to the manufacturer’s protocol; briefly nasopharyngeal swabs were placed in extraction solution, swirled 10 times and squeezed against the collection tube wall. Extracted sample was applied on a cassette with an appropriate time allowed for a monoclonal anti-SARS-CoV-2 antibody reaction. The required duration for the test to be interpreted is between15-30-min depending on the test manufacturer. There are two lines on the cassette, control line (C) which should always appear as a colored line after adding a proper sample volume. Positive result was defined by clear colored intense band at the T (test) mark on the cassette, weak positive was defined by faint to moderately intense band. Negative results indicate no visible band. If control reaction failed the test was considered invalid test and repeated. The results were read by two independent observers.

### RT-qPCR assay

RNA was extracted from viral transport media using magnetic bead NuActor Automatic Extractor (Boditech Med, South Korea). Patient samples were also analyzed by RT-qPCR within 24 hrs after collection to determine false negatives and positives (Xpert Xpress SARS-CoV-2 GeneXpert or DAAN GENE RT-PCR COVID-19 detection kit). Both assays target Nucleocapsid (N) gene for which cycle threshold (Ct) was considered in this study.

We had also analyzed 31 samples out of the total number of 231 to evaluate the reliability of re-using swabs after rapid Ag testing for RT-qPCR. There is an additional step in rapid Ag testing in which swabs are placed in lysis buffer before being transported into viral transport media (VTM). Results were compared to those obtained from swabs used directly for standard RT-qPCR procedure.

### Statistical analysis

Sensitivity, specificity, positive and negative predictive values and accuracy were calculated using MedCalc online statistical software ^6^. Laboratory COVID-19 prevalence data were obtained from Biotechnology Research Center case registry for the study period (unpublished).

Sensitivity was calculated as: (true positives)/ (true positives + false negatives) x 100 Specificity was calculated as: (true negatives)/ (true negatives + false positives) x 100

Positive predictive value:

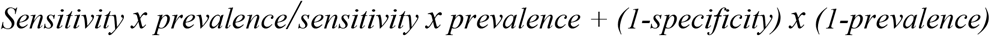

Negative predictive value:

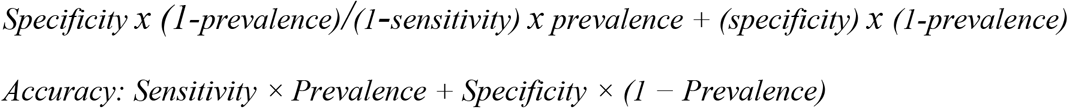

## Results

A total of 108 nasopharyngeal samples tested positive for SARS-CoV-2 by RT-PCR accounting for 46.75% of cases. The mean duration from symptom onset was 6.6 ±4.3. In this study 161 cases had symptoms consistent with COVID-19. The median cycle threshold (Ct) of positive samples was 25. Among the 108 positive samples detected by RT-qPCR, the COVID-19 Ag tests detected 83 cases correctly. Among negative samples, the COVID-19 Ag test detected all 123 samples as negative. All rapid Ag test devices used in this study showed 100% specificity. While tests from 6 manufacturers had an overall sensitivity range from 75-100%, the remaining 4 tests had sensitivity of 50-71.43% (Table 2). False negative samples had a median Ct of 34 and average duration of onset of symptoms of 11.3 days (range=5-20 days).

**Table 2.** Diagnostic performance of SARS-COV-2 rapid antigen detection tests.

| Type of antigen<br>(No. cases) | ^Detected<br>by RT-qPCR<br>(%) | ~ Not-detected by<br>RT-qPCR<br>(%) | False<br>negatives | Median Ct<br>(Range) | Symptom's<br>duration<br>Days<br>Mean ±SD | Sensitivity %<br>(Confidence<br>interval) | Specificity %<br>(Confidence<br>interval) | *PPV<br>% | ^ NPV% | Accuracy<br>% |
| --- | --- | --- | --- | --- | --- | --- | --- | --- | --- | --- |
| **FLUORECARE (21)** | 11 (52.4) | 9 (47.6) | 1 | 27.5 (21-34) | 5.3±3.3 | 91.67 (61.52 - 99.79) | 100 | 100 | 97.01 | 97.75 |
| **ESPLINE (17)** | 8 (47) | 7 (53) | 2 | 29.5 (20-37) | 6.8±3.5 | 80.00 (44.39- 97.48) | 100 | 100 | 93.11 | 94.6 |
| **RAPIGEN (39)** | 10 (25.6) | 23(74.4) | 6 | 25 (20-36) | 6.4±3.1 | 62.50 (35.4- 84.80) | 100 | 100 | 87.82 | 89.88 |
| **ASSUT (31)** | 5 (16.1) | 24 (84) | 2 | 21 (15-29) | 5±3 | 71.43 (29.04- 96.33) | 100 | 100 | 90.44 | 92.29 |
| **Orient GENE (22)** | 5 (23) | 12 (77.3) | 5 | 31 (16-39) | 7.3±4 | 50.00 (18.71- 81.29) | 100 | 100 | 84.39 | 86.5 |
| **AMP (15)** | 6 (40) | 8 (60) | 1 | 24 (18-37) | 6.3±3.2 | 85.71 (42.13- 99.64) | 100 | 100 | 94.98 | 96.14 |
| **ACON (25)** | 15 (52.4) | 10 (47.6) | 0 | 23 (18-37) | 6.7±4.4 | 100.00 (78.20- 100) | 100 | 100 | 100 | 100 |
| **ABBOTT (23)** | 10 (43.48) | 10 (56.5) | 3 | 29 (13-38) | 9.6±7 | 76.92 (46.19- 94.96) | 100 | 100 | 91.61 | 93.45 |
| **CERTEST BIOTEC (17)** | 5 (29.4) | 9 (70.6) | 3 | 22.7 (19-35) | 7±5 | 62.50 (24.49- 91.48) | 100 | 100 | 87.82 | 89.88 |
| **BIOPERFECTUS (21)** | 8 (38.1) | 11 (62) | 2 | 21 (14-34) | 6.3±4.5 | 80.00 (44.39-97.48) | 100 | 100 | 93.11 | 94.6 |

\*PPV is Positive Predictive Value, ^ NPV is Negative Predictive Value, ^ Detected by RT-qPCR denotes true positives and ~not-detected by RT-qPCR denotes true negatives.

The prevalence of positive samples calculated from Biotechnology Research Center lab database during the study period was 27%. At this prevalence rate, estimated positive predictive values were 100% and the negative predictive value ranged between 84.39%-100% for the different rapid antigen test devices.

We evaluated the sensitivity of each antigen test from the first day of symptoms up to 6 days, >7, 7-9, 10-12 and >12 days (Figure 1). Sensitivity during the first 6 days of symptoms and in samples with high viral loads (Ct<25), was 100% in all but 2 (Assut and AMP) of the test platforms (Table 3). The median RT-qPCR cycle threshold value of positive samples was 25. The majority of antigen test platforms had excellent performance during the first 6 days of symptoms with sensitivity ranging between 75-100% (Table 3).

**Table 3.**
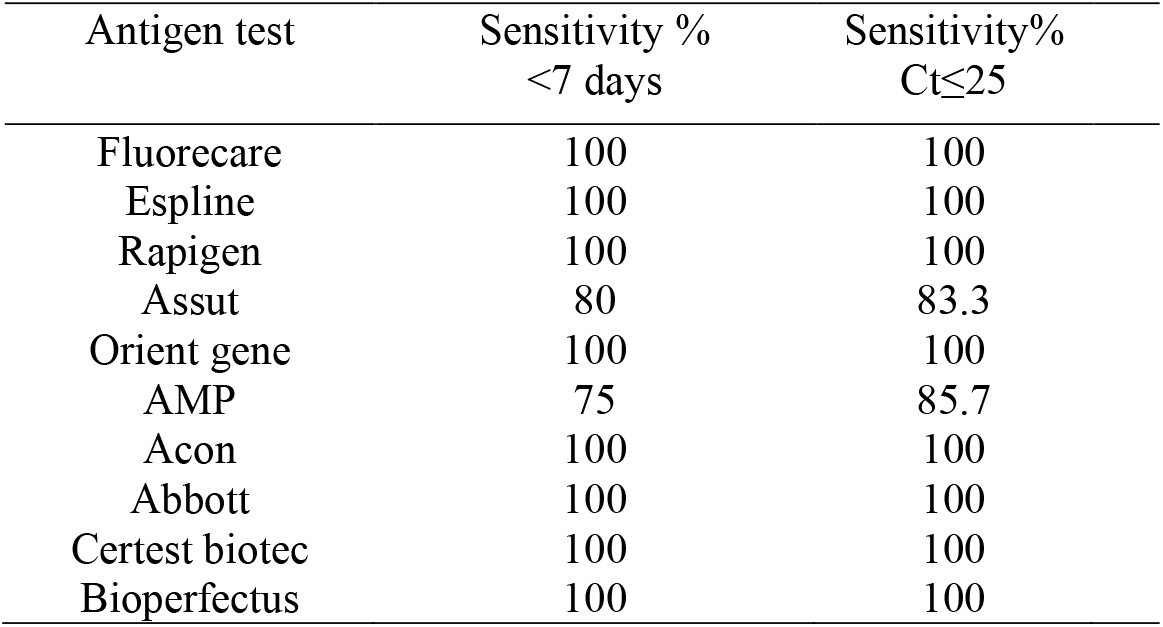
Diagnostic performance of SARS-COV-2 rapid antigen detection tests according to symptoms duration and cycle threshold.

**Fig 1.**
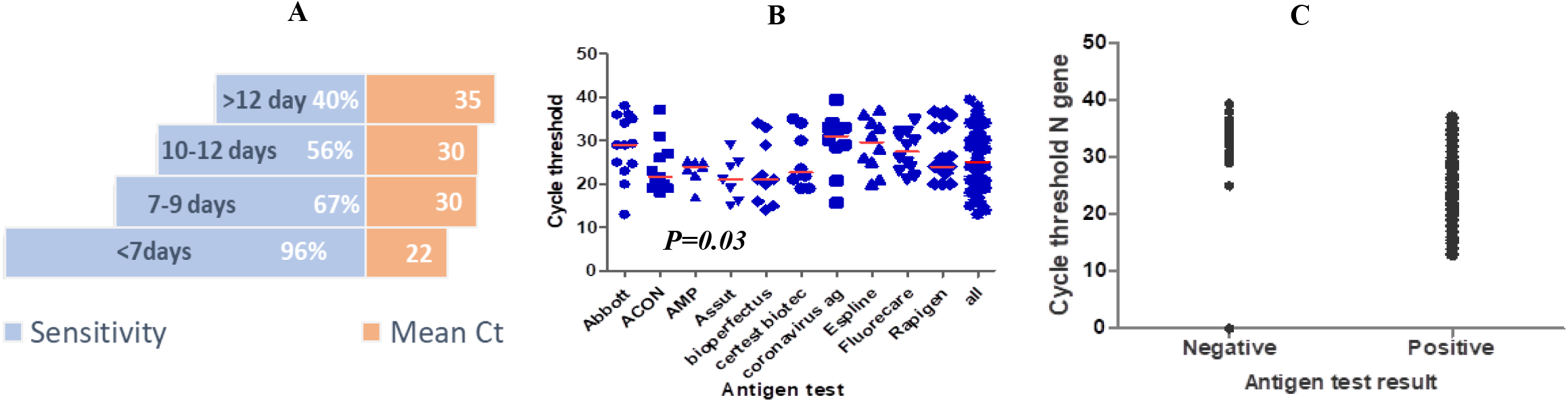
**A.** Mirror chart showing % sensitivity of rapid antigen tests by symptoms onset against corresponding mean Ct. **B** Scatter plot of Ct values of samples in each commercial test (red line =Median). **C**. SARS-CoV-2 antigen test results according to viral load.

It is worth mentioning that three participants (3/58, 5.2%) who were asymptomatic at the time of testing with history of close contact with a confirmed SARS-CoV-2 case tested positive by antigen test (Espline & Rapigen rapid Ag tests). The Cts of their RT-qPCR assay were 23, 24 and 25 and duration since contact were 5,6 and 10 days respectively. All other asymptomatic cases with exposure were correctly diagnosed as negative. All asymptomatic cases with no history of exposure were also tested negative by Ag test. Among symptomatic patients, the sensitivity was 77%.

Patients who experienced a change in smell/taste were 41. More than third of them (15/41, 36.5%) were not detected by rapid Ag tests; Abbott (no=3, mean Ct=36), Assut (no=1, Ct=29), Bioperfectus (no=1, Ct=33), CerTest (no=2, mean Ct=34.5), Orient Gene (no=3, mean Ct=32.3), Espline (no=1, Ct=36), and Rapigen (no=4, mean Ct=35). In this subgroup, the mean duration since symptoms onset was 12 days and the mean Ct was 34. However, those who had altered smell/taste and tested positive had a mean of 6.6 days since symptoms onset and mean Ct was 25. Loss of smell/taste was the only symptom in few patients (3/41, 7.3%).

Although the recommended time for interpretation ranges between 10-30 minutes, in all samples with CT< 25 the test Line was clearly visible within 2 minutes of application which gave a rough estimation of viral load. When interpretating results after or before the time set by the manufacturer, it was observed that positive test line either faded or became unreadable in most cassette assays. Also, the result interpretation before the end of recommended time was unreliable in cases with low viral load (Ct above 25).

The cycle threshold for nasopharyngeal swab positive samples used for antigen testing and then transferred to viral transport media for RT-qPCR was increased 3-6 cycles in comparison with other direct swabs.

## Discussion

Rapid antigen testing is cost-effective, easy to use and can be manufactured in large quantities. The timeliness of results they provided will reduce the load on the diagnostic laboratories. The FDA has authorized 14 SARS-CoV-2 antigen diagnostic tests for emergency use as of 19^th^ March, 2021 ^7^. Most of these tests are lateral flow assays that can be visually read. We aimed to study rapid antigen tests from different manufacturers to evaluate their performance using time since the onset of symptoms as criteria for testing. This study was also part of pre-implementation evaluation to confirm performance of the tests by RT-qPCR.

Our study showed that most Ag rapid tests examined were reliable to diagnose SARS-CoV-2 infection. They demonstrated excellent performance in samples with high viral load Ct≤25 which are usually samples taken within the first six days after the onset of symptoms. CerTest Biotech, Panbio, Rapigen, Acon Ag rapid tests demonstrated similar results to studies reported by other authors during the first week ^8–10^. The World health organization recommended the use of SARS-CoV-2 Ag rapid tests that meet a minimum sensitivity of ≥80% and ≥97% specificity compared to a nucleic acid amplification test reference assay ^2^. All tests showed a specificity of 100 % and sensitivity over 90 % for high viral load samples, with excellent levels of agreement with PCR in 8 out of 10 tests (Fluorecare, Espline, Rapigen, Orient Gene, Acon, Abbott, CerTest Biotec and Bioperfectus). Two rapid ag tests showed sensitivity below the WHO recommended value namely, Assut and AMP; 80%, and 75% respectively.

It is worth emphasizing that interpretation time should not subceed nor exceed the manufacturer’s recommended time to avoid false interpretation of the test result. The intensity of test line color is proportionally correlated with viral load. The test line in low viral load samples may appear weak or faint which may not be clearly visible to the reader. Therefore, an additional independent reader is recommended when low viral loads are expected to reduce subjectivity and confirm diagnosis but for most cases one reader is sufficient for interpreting the results. We also don’t recommend placing the same nasopharyngeal swab used for antigen test in viral transport media for RT-qPCR analysis as the viral material will be diluted and may give false negative results.

Since SARS-CoV-2 cases have been increasing with emergence of new variants^11^, and numerous antigen tests were manufactured, it is important to evaluate them before implementation. Although there is more risk of developing false negative results after 7 days of symptom onset, this is largely counter-balanced by the rapidity of the test especially when used in targeted population. Our study shows that all COVID-19 Ag tests had a good specificity for SARS-CoV-2 detection in nasopharyngeal swab samples but had a good sensitivity only for cases within 7 days of developing symptoms (higher viral loads).

We proposed a strategy for use of rapid antigen tests in symptomatic patients according to our results and experience as shown in the flowcharts (Fig 2a and b). Although the procedure is relatively easy to perform, testing with Ag rapid tests should be conducted by trained operators following manufacturer’s instructions. Interpretation of the results of rapid Ag tests should consider prevalence of the disease in the community because positive and negative predictive value of any diagnostic test is affected by disease prevalence in the population tested ^2^.

**Fig 2.A.**
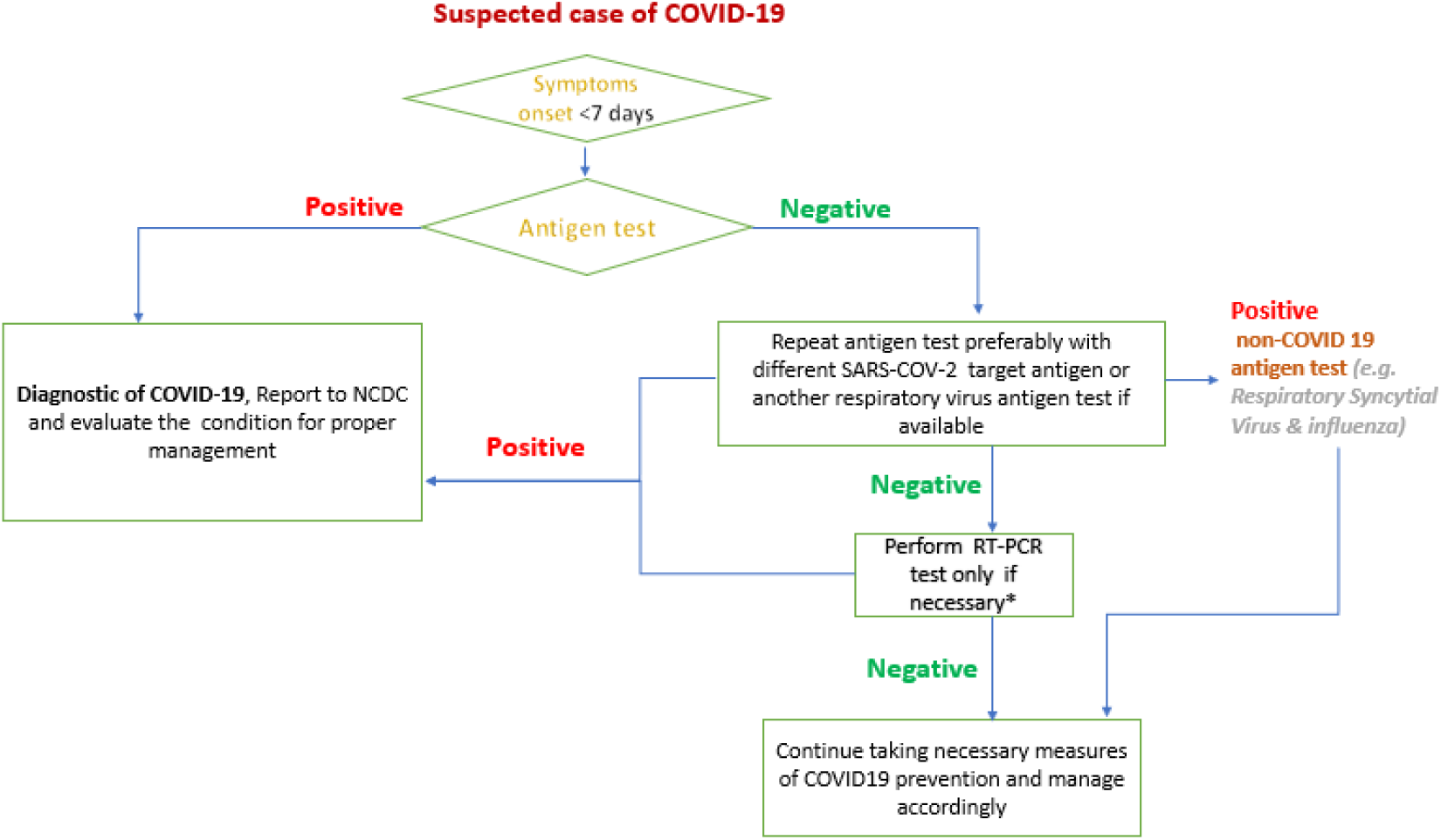
Flowchart of proposed use of rapid antigen tests in suspected COVID-19 cases with symptoms duration less than 7 days. * Vulnerable cases and elderly.

**Fig 2.B.**
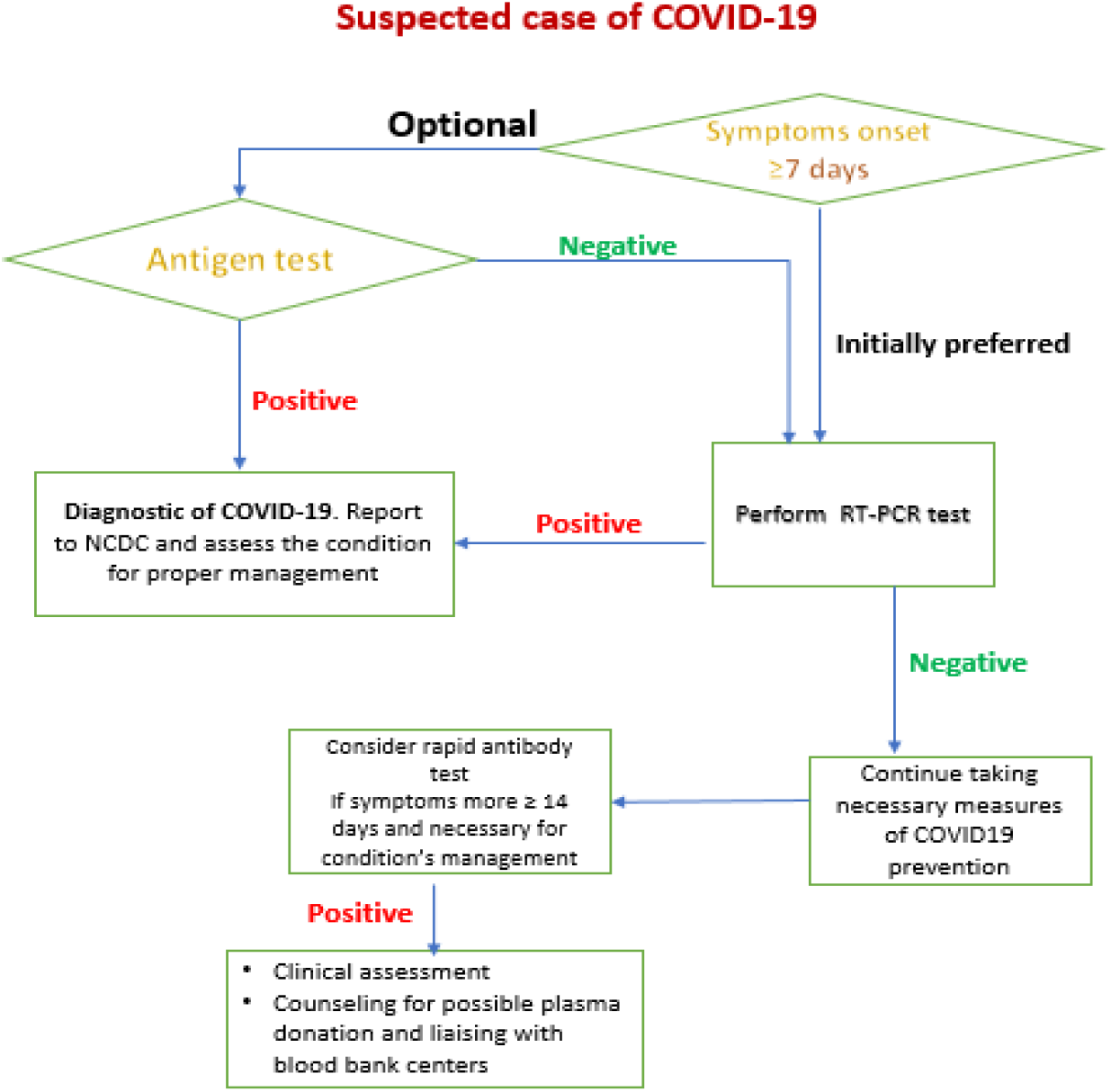
Flowchart of proposed use of rapid antigen tests in suspected COVID-19 cases with symptoms duration of more than 7 days.

In our medical departments, triage and isolation centers, we have trained medical staff on how to use the antigen test properly, developed criteria for testing (figure 2) and audited the procedure. It is important to note that the use of rapid antigen test requires careful respiratory sample management, waste disposal and biosafety considerations.

The incubation period of SARS-CoV-2 infection was estimated between 3 and 6 days with a median incubation of 4 days ^12,13^. Patients may only present to the hospital 6-10 days following symptom onset ^14^, therefore, missing the opportunity for rapid detection using antigen tests. We consequently emphasize the importance of encouraging patients to seek medical advice and immediate testing as soon as they develop any COVID-19 related symptoms. In particular when patients lose sense of smell, they should be tested immediately, preferably within 24 hrs. because this symptom usually occurs after onset of other COVID-19 symptoms ^15,16^. We also observed in our study that most patients present with loss of smell few days after symptoms onset and in some patients, it was the sole symptom (12 patients with loss of smell came late). We believe that Ag testing would encourage patients to come earlier for testing because of reduction in waiting time.

Viral culture studies in cell lines showed that samples with Ct value ≥ 34, ≥ 24 or ≥ 38 and more than 8 days of symptoms onset had no growth and thus might indicate the person is less infectious. ^17–19^. Thus, there is no agreement about the cut-off Ct value. A positive PCR results reflects only the detection of viral RNA and not necessarily indicate the presence of viable virus. Therefore, false negative results could potentially be non-infectious, increasing the safety margin in the utility of rapid antigen test in cases more than 7 days after start of symptoms and asymptomatic patients.

This study has some limitations. First, the sample size was small for each individual antigen rapid test kit. Second, because the data was collected prospectively based on the presence of symptoms or history of contact with an infected patient, it was difficult to select patients with different Ct values in enough size. However, if we assumed that the performance of all antigen rapid test devices was similar, the data would be reliable enough to give credible results.

Rapid Ag testing performance is mostly affected by symptoms duration, viral load, manufacturing company, operator experience and qualification. In addition, new SARS-COV-2 variants may alter the performance especially with spike proteins targeted rapid ag tests ^20,21^.

In conclusion, rapid Ag tests have high sensitivity and specificity in early disease when patients present before 7 days of symptom onset. Patients are encouraged to test as soon as they get COVID-19 related symptoms within 1 week and to seek medical advice within 24 hrs. if they develop disturbed smell/taste. The use of rapid antigen tests is important for controlling COVID-19 pandemic and reducing burden on molecular diagnostic laboratories.

## Data Availability

Data will be available upon request

## Acknowledgment

The authors gratefully acknowledge the support of the Libyan authority for research, science and technology and all lab researchers at Biotechnology Research Center COVID-19 detection team.

## Author contribution statement

The corresponding authors had full access to all the data in the study and have final responsibility for the decision to submit for publication. ZA and IA contributed equally and share first authorship. Concept and design: ZA and IA. Acquisition, analysis, or interpretation of data: ZA, IA, HK, SE and FE. Drafting and revising the manuscript: ZA, IA. KI, TD. Statistical analysis: TD, AE and MB. Administrative, technical, or material support: SE, HK and FE. Supervision: SA, AE.

## Ethical approval

Ethical approval All procedures performed in studies involving human participants were in accordance with the ethical standards of the institutional and/or national research committee and with the 1964 Helsinki declaration and its later amendments or comparable ethical standards. This study was approved by the Biotechnology Research Center Bioethics Committee.

## Informed consent

Informed consent was obtained from all individual participants included in the study.

## Data Availability

Data available on request from the authors. The data that support the findings of this study are available from the corresponding author upon reasonable request.

